# Prevalence mapping of Schistosomiasis among Pre-school aged children in Rwanda

**DOI:** 10.1101/2022.01.26.22269869

**Authors:** Bayingana Jean Bosco, Nyandwi Elias, Ntakarutimana Amans, Kagabo Joseph, Shema Eliah, Kanimba Philbert, Mbonigaba Jean Bosco, Ruberanziza Eugene, Rujeni Nadine

## Abstract

Schistosomiasis is endemic in Rwanda and control programs have been implemented with a special focus on school-aged children (SAC) ignoring pre-school aged children (pre-SAC) for which the actual prevalence of the disease is not well established. This study consisted of a cross-sectional quantitative mapping of the distribution of schistosomiasis and identification of associated risk factors among pre-SAC throughout the country. The study covered all the 17 Districts of Rwanda endemic for schistosomiasis with a total sample of 4675 children enrolled from 80 purposively selected villages. The Parasitological assessment of children’s urine and stool samples was conducted using CCA and Kato Katz methods respectively for infection detection. A standard questionnaire was used to collect data on the risk factors and geospatial assessment was performed using tablets and GPS to record geographic coordinates for plotting locations on maps using ArcGIS software.

The overall prevalence of *S mansoni* infection across the surveyed areas was 24% and 0.8 by CCA and Kato-Katz, respectively. Infection was significantly associated with bathing children in open water bodies. Furthermore, pre-SAC looked after by siblings (sisters) were twice as much likely to be infected compared to those looked after by mothers. Schistosomiasis control interventions are needed for pre-SAC to limit their exposure to open water bodies with expectations of adapted chemotherapy to be availed. Community based deworming campaigns may be the best way to ensure good treatment coverage of pre-SAC in Rwanda.

**Author summary:** Schistosomiasis is one of the Neglected Tropical Diseases (NTD) of public health concern in Rwanda like in many tropical countries. The recently published NTD roadmap by the World Health Organization indicates that schistosomiasis is targeted for elimination as a public health problem worldwide by 2030. For this target to be achieved, all at risk populations should be reached by control programs’ interventions such as preventive chemotherapy, health education as well as water, sanitation and hygiene (WASH) practices. However, pre-school children (pre-SAC) are among populations at risk for whom infection burden is not fully documented. Implementation of the recent WHO guideline on schistosomiasis control and elimination that recommend inclusion of pre-SAC in control programs will be informed by detailed assessment of the infection burden and distribution in endemic countries. This study is showing for the first time a high burden of schistosomiasis among pre-SAC in high-risk areas of Rwanda. With the imminent availability of the paediatric formulation of Praziquantel (the drug of choice against the disease), the findings should guide the country in implementation strategies that include these children in mass deworming. The findings also highlight a number of risk factors including the passive exposure of these young children by their siblings while bathing them in open waters and the lack of knowledge of parents/guardians on the disease. Additional integrated interventions such as health education, improvement of water supply and sanitation as well as snail surveillance will lead to more sustainable solutions in the march towards schistosomiasis elimination.

## Introduction

Schistosomiasis is a chronic parasitic infection that is caused by trematodes of the genus *Schistosoma*, transmitted through fresh water snails. It can cause vascular damage in individuals and is associated with growth retardation and impaired metabolism and cognition (1,2). Schistosomiasis remains a public health concern and highly prevalent in low and middle-income countries, including Rwanda, mostly affecting poor communities with no access to potable water (3). Globally, more than 250 million people are affected by schistosomiasis and Sub Saharan Africa accounts for 90% of all these infection burdens (4,5). Control interventions mainly rely on regular targeted treatments (using Praziquantel) of populations in endemic areas coupled with health education. Although the disease is targeted (as per the WHO roadmap) for elimination as a public health problem (defined as prevalence of <1% of heavy intensity schistosomiasis infections) by 2030 (6), control interventions do not currently reach all populations in need in most endemic areas.

For many years, pre-school aged children (pre-SAC) have been left out in most schistosomiasis treatment programs. This was partly due to the assumption that they are not at great risk of contracting schistosomiasis. However, considerable advances have been made in this regard and a number of studies have shown that most of the pre-SAC in endemic areas are significantly exposed to schistosomiasis in many countries (7–14). This has prompted the WHO to formulate a new guideline including recommendations to include pre-SAC aged 2 years+ in preventive chemotherapy while those below 2 years should be treated on individual basis (15).

With the realization that pre-SAC are in need of treatment, came a number of challenges including the absence of an appropriate child friendly formulation of Praziquantel and the lack of information on the real burden of schistosomiasis in this young age group in some countries. Indeed, inclusion of pre-SAC in schistosomiasis control programs should be justified by quantification of the burden of infection in that age group in each country for optimal intervention. Furthermore, an assessment of their accessibility and local acceptability should guide implementation strategies.

Rwanda is among the countries where the burden of schistosomiasis among pre-SAC is not well documented. Our own study on Nkombo Island in the western part of the country reported an overall prevalence of 9.5% (9), but a countrywide assessment of the disease in this age group is necessary to better inform the control program. Nevertheless, the Rwandan Ministry of health has recently introduced a community-based deworming strategy whereby community health workers (CHWs) and community leaders coordinate MDAs with minimal supervision from the central level. Such a deworming strategy should be suitable for treating schistosomiasis among pre-SAC.

In light of the above, this study was conducted to estimate the prevalence, identify risk factors and produce an illustrative, countrywide map of schistosomiasis distribution in all moderate to high - risk areas of the country.

## Methods

### Study design, population, and sample size

This was a cross-sectional quantitative study that involved parasitological assessment of schistosomiasis among children aged 7 to 59 months. Structured interviews were also held with participants’ parents/caregivers to assess risk factors of infection in this young population. The study covered all the 17 Districts of Rwanda at moderate to high risk of schistosomiasis (16,17) and targeted 40 Sectors reported with a prevalence of 10% and above among school children (based on incidence and mapping data). From each targeted sector, 2 villages were selected based on their proximity to existing water body and/or wetlands. The villages layer was overlaid with layers of lakes, multipurpose water dam, fish pond and/or important wetlands (wetland of ≥ 0.7 ha hosting a socio-economic activity, mostly irrigated agriculture). From eligible villages (risky villages), final selection was purposively decided considering the spatial distribution pattern analysed visually by researchers and complemented with field verification, making 80 study villages with proximity to open water sources and wetlands. A total of 4675 children were enrolled from the selected villages.

### Parasitological assessment

Urine samples were collected and tested for schistosomiasis circulating antigen using the Point-Of-Care Circulating Cathodic Antigen (POC-CCA or CCA) following manufacturer’s instructions. CCA test results were recorded as negative (- or trace) or positive (+; ++ or +++ according to the intensity of the test line in comparison to a test control). Stool samples were also collected and tested using the Kato Katz method for the detection of patient infection following published protocols (18). A single specimen was collected from which 2 slides were prepared and read by 2 laboratory technicians independently. Ten per cent (10%) of all slides were re-tested by the National Reference Laboratory senior technicians for quality control.

### Risk factors and spatial assessment

A standard questionnaire was designed in Kobo toolbox and used to collect data from parents/guardians by a well-trained data collector using a tablet. Geographic coordinates were recorded using tablets and ArcGIS software version 10.4 was used to produce maps.

### Statistical analysis

Data collected were analysed by Stata 13 (STATA Corp, Lakeway, College Station, Texas, USA). Descriptive analysis was conducted. Categorical variables were presented as frequencies and their respective percentages. Numerical variables were presented as mean and standard deviations. For comparison purposes, cross-tabulation analyses were conducted and contingency tables were produced, while chi-square tests were used to compare proportions. To assess the association between variables, logistic regression was performed. For binary logistic regression, each risk factor was assessed with the outcome (infection status) separately while for multivariable logistic regression, a backward stepwise logistic regression was run and all risk factors with a p-value less or equal to 0.05 were considered. Odds ratios and the corresponding 95% confidence interval (CI) were reported. A two-tailed significance level of 0.05 was considered.

### Ethical consideration

The current study was approved by the Institutional Review Board (IRB) of the University of Rwanda College of Medicine and Health Sciences (Approval Notice No 148/CMHS/IRB/2019) and endorsed by the Ministry of Health. Local authorities, community health workers and parents/guardians were informed about the aims, voluntary nature of their participation, potential risks and benefits of the study. A signed consent was obtained from parents/guardians before enrolling their child(ren) in the study.

## Results

The results of this research are presented by study sites and do not represent the administrative District. Study sites are those sites in proximity with open water bodies and vary in number per District from one to six. These sites are the base for defining the targeted areas of control interventions.

### Demographic characteristics of the study participants

The total sample was 4675 pre-SAC with a mean age of 37.5 months (±13.0), and a sex ratio M:F of 1.01: 1 (Table 1). Majority of the study participants (corresponding to 96.2%) were born in their respective villages (permanent residents).

**Table 1.**
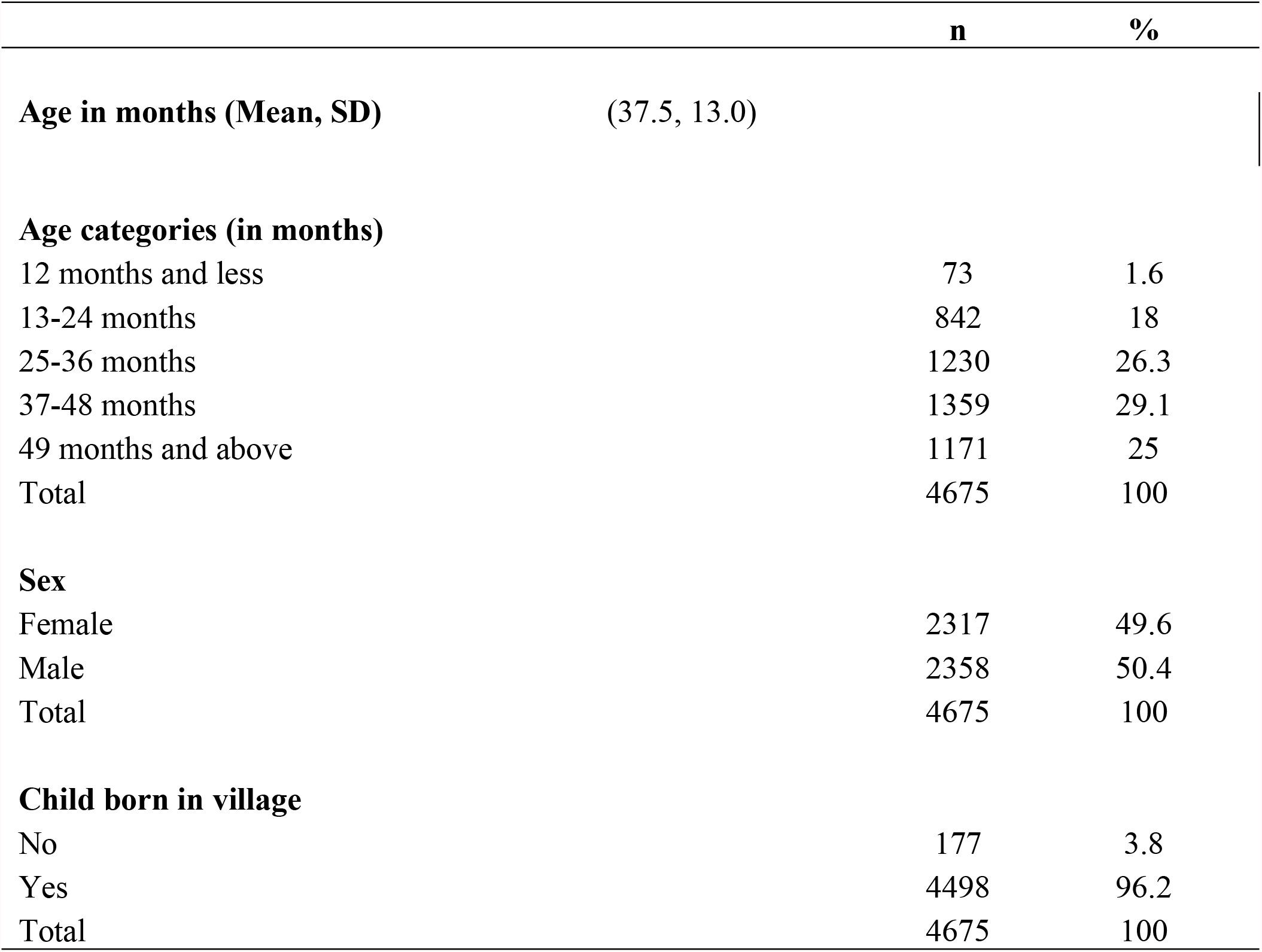
Characteristics of pre-school aged children enrolled in the study

### Water contact activities (exposure) for children in study areas

More than 90% parents reported farming as their occupation and 83.3% (n= 3895) of the families use open water sources (lake, dam or swamp) for daily domestic activities. This result in more than 60% of enrolled children being regularly in contact with open water sources (Table 2).

**Table 2.** Water contact activities for parents and their children in the study areas

|  | **n** | **%** |
| --- | --- | --- |
| **Parent's occupation** |  |  |
| Casual worker | 143 | 3.18 |
| Farmer | 4,060 | 90.26 |
| Government employee | 52 | 1.16 |
| Self employed | 157 | 3.49 |
| Unemployed | 86 | 1.91 |
| Total | 4,498 | 100 |
| **Fetching water in the open water source** |  |  |
| No | 780 | 16.7 |
| Yes | 3895 | 83.3 |
| Total | 4675 | 100 |
| **Child taken to open water source** |  |  |
| No | 1444 | 30.9 |
| Yes | 3222 | 69.1 |
| Total | 4666 | 100 |
| **Frequency child taken to lake** |  |  |
| Once a day | 2055 | 63.4 |
| Once a week | 824 | 25.4 |
| Once a month | 185 | 5.7 |
| Once a year | 53 | 1.6 |
| Once in his life | 123 | 3.8 |
| Total | 3240 | 100 |
| **Child bathed in the lake** |  |  |
| No | 1669 | 36.2 |
| Yes | 2937 | 63.8 |
| Total | 4606 | 100 |
| **Frequency child bathed IN the lake** |  |  |
| Once a day | 1943 | 65.2 |
| Once a week | 761 | 25.5 |
| Once a month | 155 | 5.2 |
| Once a year | 38 | 1.3 |
| Once in his life | 83 | 2.8 |
| Total | 2980 | 100 |
| **Child ever bathed using FETCHED lake water** |  |  |
| No | 947 | 20.3 |
| Yes | 3728 | 79.7 |
| Total | 4675 | 100 |

### Prevalence and infection intensity of schistosomiasis among pre-school aged children

Prevalence of schistosomiasis among pre-school aged children was assessed using both KK and POC-CCA. The POC-CCA results yielded a prevalence of 24% among children tested (trace results considered negative), while the KK only yielded a 0.8% prevalence. The POC-CCA estimate was considered as the overall prevalence given the high sensitivity of the assay (16). For infection intensity, KK results were considered and the raw faecal worm egg count (FWEC) was multiplied by 24 to estimate the eggs per gram (epg) (18,19). Based on WHO guidelines (20), 16.2% (n= 6) of the 37 children who tested positive on Kato Katz had moderate infection while the rest had light infection (Table 3).

**Table 3.**
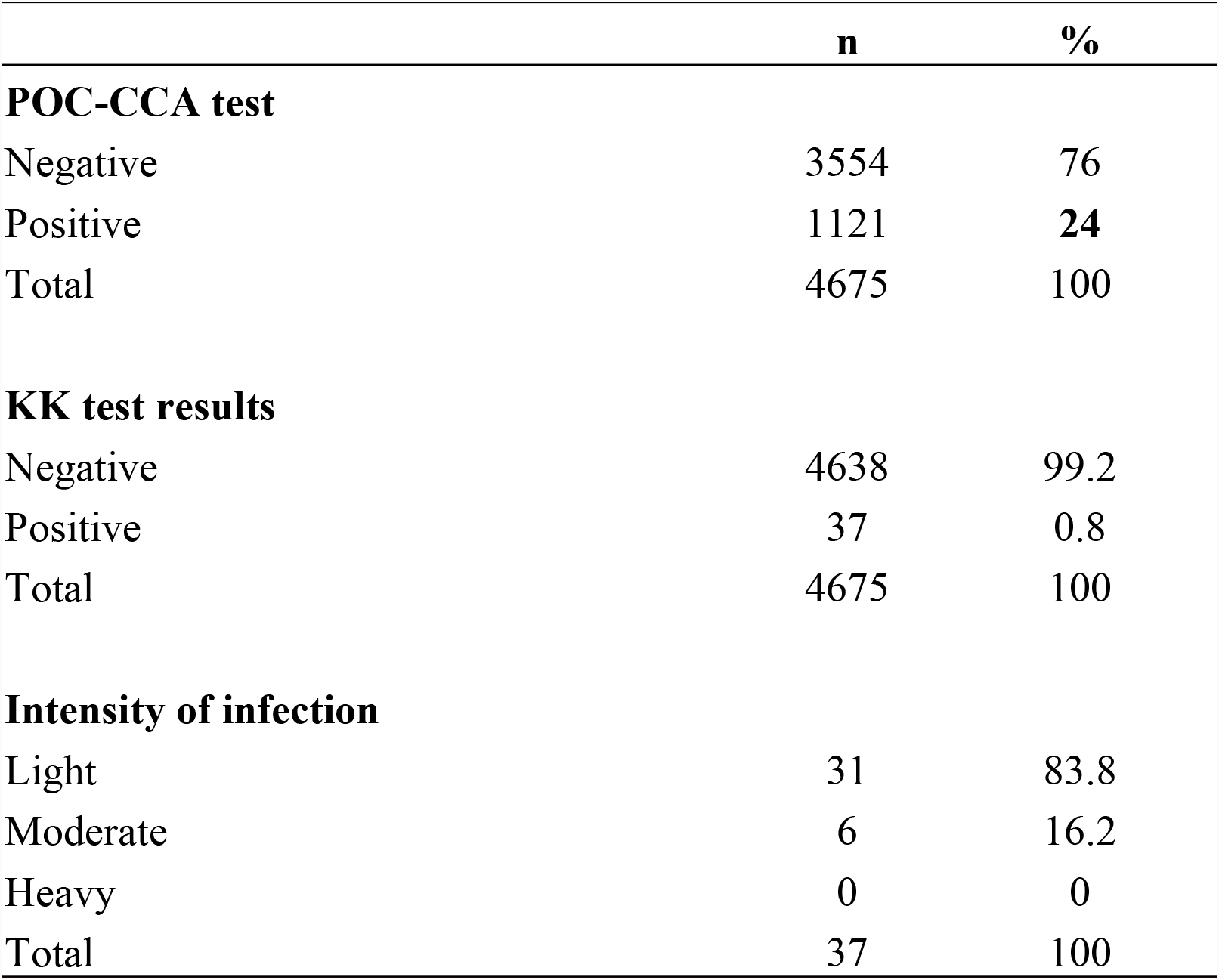
Prevalence and infection intensity of the study population

|  | **n** | **%** |
| --- | --- | --- |
| **POC-CCA test** |  |  |
| Negative | 3554 | 76 |
| Positive | 1121 | **24** |
| Total | 4675 | 100 |
| **KK test results** |  |  |
| Negative | 4638 | 99.2 |
| Positive | 37 | 0.8 |
| Total | 4675 | 100 |
| **Intensity of infection** |  |  |
| Light | 31 | 83.8 |
| Moderate | 6 | 16.2 |
| Heavy | 0 | 0 |
| Total | 37 | 100 |

### Prevalence of schistosomiasis by age, gender, and location among pre-school children

The results on the prevalence of schistosomiasis infection by gender revealed no statistically significant difference in infection between males and females. However, a statistically significant difference in infection was observed across age groups (Table 4).

**Table 4.** Infection prevalence per age category and gender.

| Age |  | CCA – traces considered negative |  | KK |  |
| --- | --- | --- | --- | --- | --- |
|  |  | Negative | Positive | Negative | Positive |
| ≤12 months | n | 52 | 21 | 73 | 0 |
|  | % | 1.5 | **1.9** | 1.6 | **0.0** |
| 13-24 months | n | 617 | 225 | 841 | 1 |
|  | % | 17.4 | **20.1** | 18.1 | **2.7** |
| 25-36 months | n | 915 | 315 | 1222 | 8 |
|  | % | 25.7 | **28.1** | 26.3 | **21.6** |
| 37-48 months | n | 1068 | 291 | 1346 | 13 |
|  | % | 30.1 | **26.0** | 29.0 | **35.1** |
| ≥ 49 months | n | 902 | 269 | 1156 | 15 |
|  | % | 25.4 | **24.0** | 24.9 | **40.5** |
| | | $\chi^2$ (P-value) = 11.7493 (**0.019**) | | $\chi^2$ (P-value) = 9.7921 (**0.044**) | |
| **Gender** |  |  |  |  |  |
| Female | n | 1782 | 535 | 2298 | 19 |
|  | % | 50.1 | 47.7 | 49.5 | 51.4 |
| Male | n | 1772 | 586 | 2340 | 18 |
|  | % | 49.9 | 52.3 | 50.5 | 48.6 |
| | | $\chi^2$ (P-value) = 1.9890 (0.158) | | $\chi^2$ (P-value) = 0.0478 (0.827) | |

There were significant differences in infection prevalence across study sites, Gisagara District having the highest prevalence with 60.4%, followed by Nyagatare and Gasabo Districts respectively with 46.8% and 40%, while Kamonyi District had the lowest prevalence (7.1%). Infection distribution across study sites is illustrated on Fig 1.

**Fig 1.**
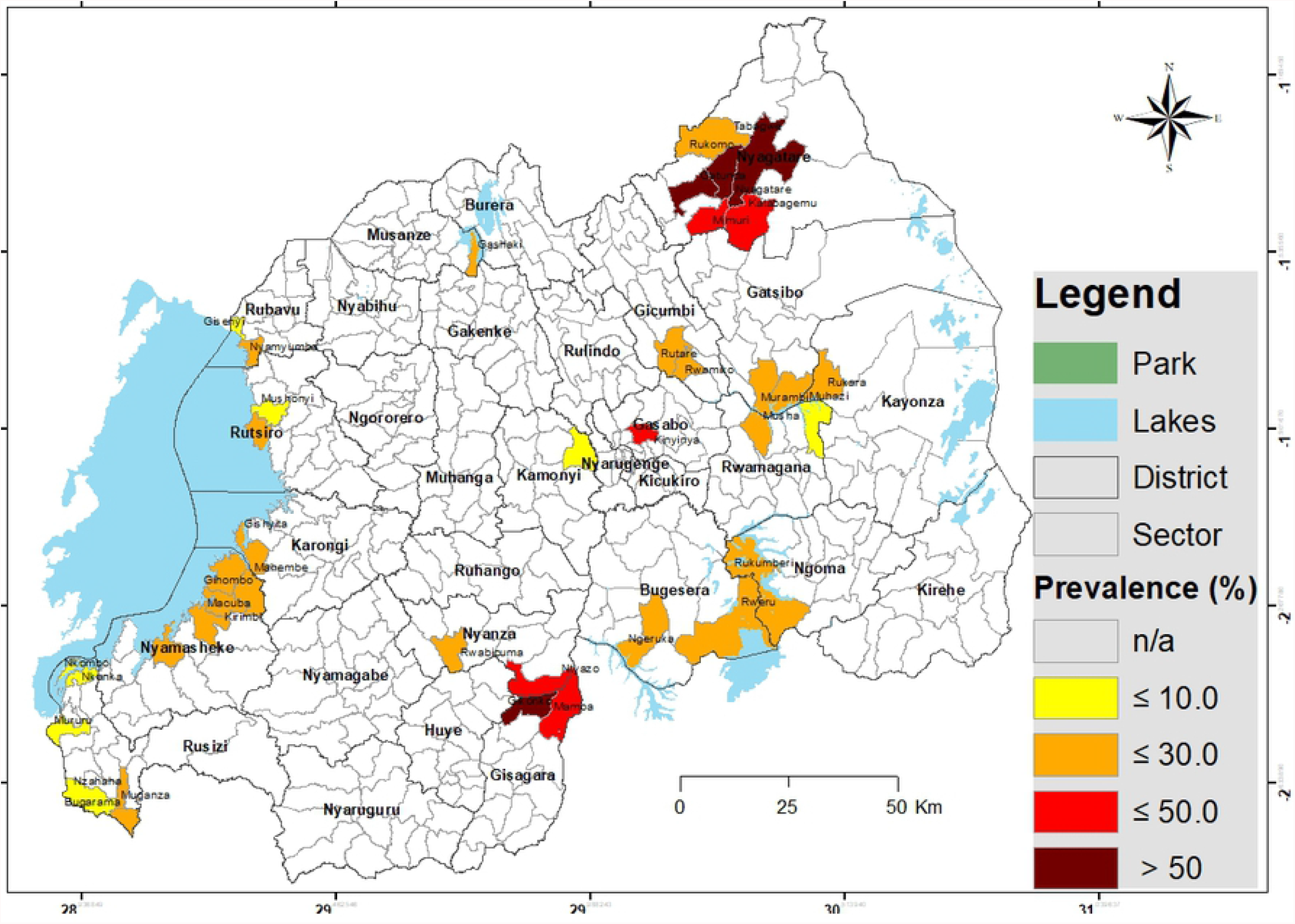
Map illustrating the distribution of schistosomiasis infection among pre-SAC in Rwanda. Infection prevalence was based on CCA findings (trace as negative); Geographical coordinates were collected for each site (village) and were extrapolated to the lowest administrative boundary (sector).

### Factors associated with Schistosomiasis infection

The logistic regression performed highlighted the age as a contributing factor to infection status as expected. In addition, two behavioural factors were significantly associated with the occurrence of infection: children taken care of by a sister and those who are bathed in the lake/pond are approximately 2 times more at risk of being infected (OR = 1.9, p<0.0001 and OR=1.8, p<0.0001 respectively) compared to their counterparts.

### Knowledge on Schistosomiasis and perception on community based deworming

Approximately 35% (n=1629) of parents/ caregivers had never heard of schistosomiasis (or Bilharzia) before. However, out of the 155 community health workers (CHWs) interviewed, 98.7% reported knowing the disease by name and 96.1% knew that it is contaminated through contact with water bodies and that pre-SAC are also at risk of contamination (Table 7).

**Table 6.**
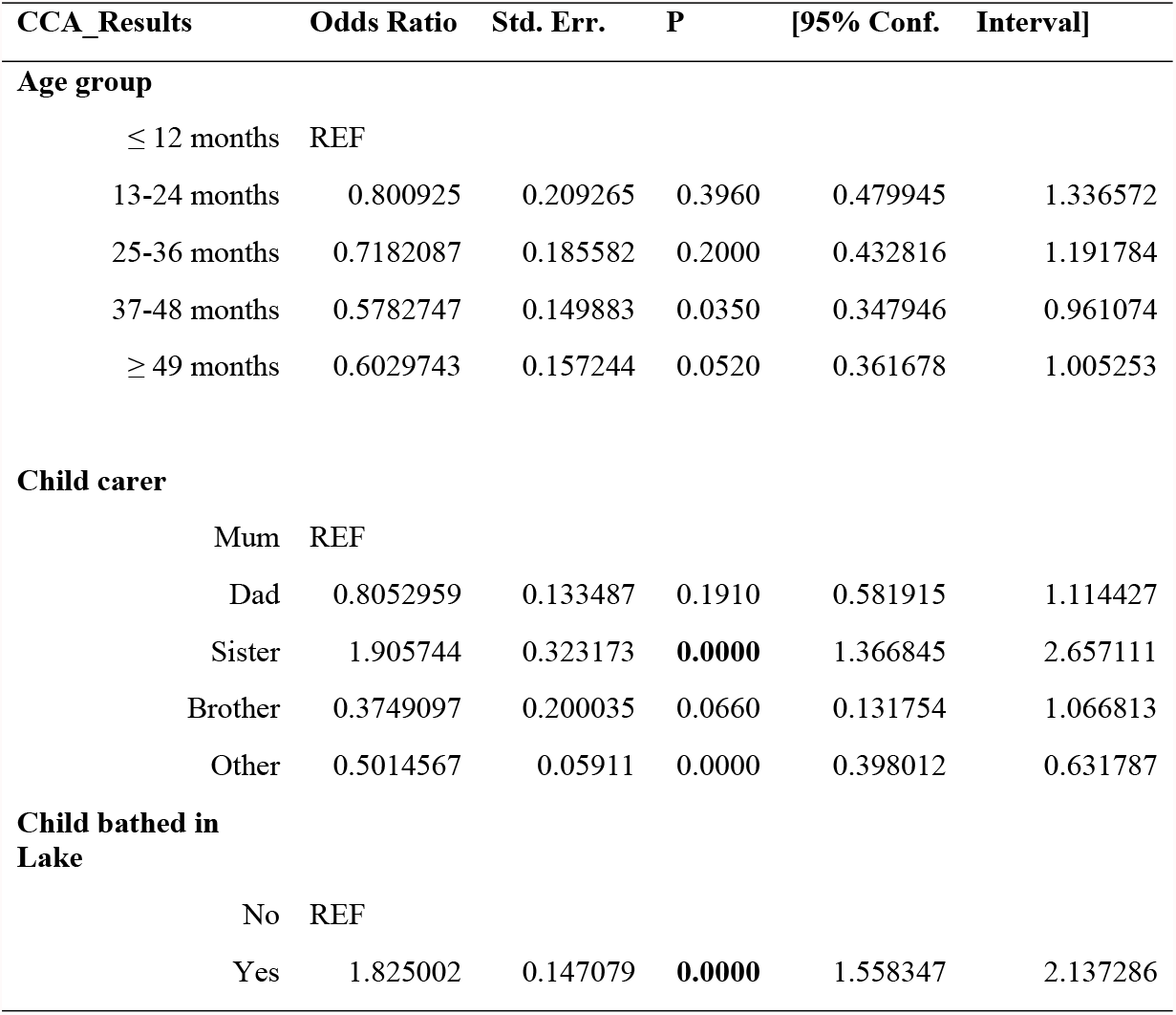
Association between behavioural factors and schistosomiasis.

**Table 7.** CHWs’ general knowledge on schistosomiasis

|  | n | % |
| --- | --- | --- |
| **Ever heard of Bilharzia** |  |  |
| No | 2 | 1.3 |
| Yes | 153 | 98.7 |
| Total | 155 | 100 |
| **How is schistosomiasis/bilharzia acquired?** |  |  |
| Don't know | 6 | 3.9 |
| Contact with water bodies | 148 | 96.1 |
| Total | 154 | 100 |
| **Can children under 5 years be infected?** |  |  |
| No | 1 | 0.6 |
| Yes | 151 | 97.4 |
| Don't Know | 3 | 1.9 |
| Total | 155 | 100 |

Regarding the implementation of a community-based mass drug administration (MDA), 62.1% (n=95) of CHWs reported that this is implemented at their respective villages, while 37.9% (n=58) reported that it is not. However, of those (n=58, i.e. 37.9%) who reported that MDA is not yet implemented at their villages, 87.9% (n=51) strongly support the idea of introducing it in their villages. Majority of the CHWs (99.3%, n=136) reported that they are willing to be involved in the implementation of community-based MDA campaigns in their village and 77.9% (n=113) suggested that the village leader would be the best person to coordinate this activity. CHWs also acknowledged some advantages of a community-based MDA such as full treatment coverage, time saving and an opportunity for health education (Table 8). However, some challenges were also highlighted such as the lack of distribution spaces, poor management of drugs and poor knowledge of CHWs at the village level.

## Discussion

The present study aimed to estimate the burden of schistosomiasis among pre-SAC in Rwanda and risk factors for the infection as well as to map the distribution of infection throughout the country for that category of population. This is the first comprehensive mapping exercise conducted in this young age group that has long been neglected by the Rwanda schistosomiasis control program. The study covered all the 17 Districts that had been highlighted by the previous school-based mapping as endemic areas (16,17). The 80 study villages as minimum units of observation (sampling units) within these endemic Districts were purposively selected for their proximity to open water bodies to allow for the focal nature of the disease distribution. From the targeted districts, the results are presented at the level of study sites which have been extrapolated to administrative sectors based on the layout of open water bodies, environmental sanitary conditions and exposed population. Overall, the prevalence of schistosomiasis was 24% using the CCA diagnostic method and 0.8% using the KK method. This discrepancy was expected as the KK has shown a low sensitivity (21), especially in pre-SAC who are most likely carrying pre-patent and/or light infections (22). Nevertheless, the CCA prevalence detected was comparable to findings among SAC in the same areas (16), highlighting an urgent need to include these children in treatment campaigns. As expected, the distribution of infection was skewed, few Sectors harbouring most of the infection burden as highlighted by the significant differences between sites (i.e 60.5%, maximum and 7.1%, minimum). In agreement with the previous mapping among SAC (16,17), sectors of Nyagatare and Gisagara Districts showed the highest infection prevalence (in 4 sectors the prevalence exceeded 50%) but also the Districts of Ngoma, Bugesera, Gatsibo, Kayonza, Nyamasheke, Gicumbi and Karongi were heavily burdened. With regard to infection intensity, and based on WHO criteria (20), most of the pre-SAC showed light to moderate infection intensities.

In this study, 83.3% of parents/guardians fetched their water for domestic use from open water sources and 63.4% confirmed taking their children to these water sites every day. Most importantly, 63.8% of the parents/guardians interviewed bath their children in open water bodies and our logistic regression analysis indicate that their children are twice as much likely to be infected compared to those who are not bathed in these waters. Previous studies elsewhere have also reported this passive nature of exposure to schistosomiasis among pre-SAC (11,23,24). Interestingly, our logistic regression highlights that children who are looked after by a sibling (a sister in this case) –most likely while their parents conduct domestic/professional chores-are more likely to be infected compared to those who are constantly with their mothers. Although we were not able to conduct a thorough observational investigation, these findings suggest that older sisters play and/or bath their younger siblings at water contact points that are far enough from where mothers wash dishes/clothes, and therefore far from detergents that may destroy cercaria (25). The latter may protect –to some extent-those children who are constantly with their mothers.

At the time of the study, 65% of the parents/caregivers had some vague information on schistosomiasis “*yes I heard about schistosomiasis*” but no extensive assessment of the level of knowledge was done. Nevertheless, more than 90% of CHWs reported that “*schistosomiasis is transmitted through open waters*”. The study was however limited in terms of depth of health education assessment as the main focus was on screening infection in the child and interviewing the caregivers to assess exposure risk factors. Nevertheless, given the findings on infection prevalence, schistosomiasis control interventions are specifically needed that focus on health education for household members’ behaviour change and practices to (a) limit the children’s exposure to open water bodies, and (b) improve children’s environmental sanitary conditions (26). Additional interventions such as provision of potable water supplies to these vulnerable communities as well as public latrines may provide longer term solutions. Indeed, more than 90% of the interviewed parents/guardians are farmers and most of our study sites were in close proximity to wetlands that are exploited for agriculture. Farmers generally spend several hours in the fields and, in the absence of latrines, are likely to defecate in these fields and contaminate the water bodies that are later used for bathing children.

The Rwandan Ministry of health has recently started implementing a community-based deworming that take place during the mother-child health (MCH) week. Our study involved interviews of CHWs to find out how they perceive this deworming approach and findings indicate that they strongly support it and highlight several advantages. This should be the best way to reach pre-SAC once the pediatric formulation of Praziquantel is availed.

Taken together, the present study findings stress that pre-SAC in Rwanda are part of the population at risk of schistosomiasis and control interventions of the disease in Rwanda must include them. In particular, we provide a detailed map of pre-SAC infection distribution that should guide intervention strategies countrywide. Further assessment of the parents/CHW knowledge, attitudes and practices towards schistosomiasis and WASH recommended practices is warranted.

## Data Availability

The data underlying the results presented in the study are available from the corresponding author

## Acknowledgements

This research was commissioned by the National Institute of Health Research (NIHR) Global Health Research programme (16/136/33) using UK aid from the UK Government. The authors would like to extend thanks to the study participants, the University of Rwanda, Rwanda Biomedical Center, Ministry of Health, district hospitals and health centers for all facilitations to the end of this research activity.

## Supporting information caption

